# Comparison of the burnout among medical residents before and during the pandemic

**DOI:** 10.1101/2022.05.28.22275707

**Authors:** Hüseyin Küçükali, Sezanur Nazlı Türkoğlu, Shams Hasanli, Fatma Nur Dayanir Çok, Hazal Cansu Culpan, Osman Hayran

## Abstract

**Objective:** This study aims to compare the level of burnout syndrome in medical residents before and during the COVID-19 pandemic and identify potential risk factors.

**Methods:** This cross-sectional study was conducted on medical residents from three different university hospitals in Turkey in March 2021, one year after the pandemic hit Turkey. Burnout is measured by the Maslach Burnout Inventory which assesses three dimensions of it: emotional exhaustion, depersonalization, and personal accomplishment. Collected data were combined and compared with data from a previous study held in the same hospitals in December 2019, three months before the pandemic.

**Results:** 412 medical residents from three universities participated. The mean age was 27.8±2.4 and half of them were female. Compared to pre-pandemic levels, no significant differences in emotional exhaustion (pre:19.0±7.6 post:18.8±7.8), depersonalization (pre:7.3±4.3 post:7.2±4.4), and personal accomplishment (pre:20.8±5.1 post:21.1±5) scores were observed one year after the pandemic. Adjusting for confounders, multiple linear regression models indicated that those who are female, are in a surgical speciality, have vulnerable cohabitants, and have more night shifts face higher emotional exhaustion. Depersonalisation is higher among those who spent more years in residency, have more night shifts, or have COVID-19 outpatient duty. Females and those who have vulnerable cohabitants have lower levels of Personal Achievement.

**Conclusion:** This study does not support the hypothesis that pandemic increases the burnout levels. Yet it identifies a couple of pandemic-related factors that are associated with burnout and confirms the association of several previously known factors.

## INTRODUCTION

Resident physicians in Türkiye are more precarious than ever. Physician identity was wounded before the pandemic (Koytak, 2021). The recent decrease in purchasing power and public disputes on physician wages did not do better. Türkiye is already one of the countries with the lowest doctor per population (1.8/1000) among OECD countries (average of 3.5) (OECD, 2021). Considering “The Great Resignation” (Sheather & Slattery, 2021) following the COVID-19 pandemic, a possible wave of resignations may intensify the problems arising from inadequate human resources, such as short consultation times and long waiting times. Because the residency program also works as a tool for distributing doctors after the training more equally among regions, this may also play a role in health inequalities. Lower completion of the residency program would mean fewer specialists in the following years. This may risk the achievements of universal health coverage in Türkiye, particularly for some specialist care. Burnout is known to be a major driver of dissatisfaction and withdrawal from job among medical doctors and studies are alarming about an increase during the pandemic (Sheather & Slattery, 2021).

Burnout has been a mental and physical syndrome characterized by emotional distress that usually occurs in individuals who need to use emotional resources as part of their work (Maslach & Jackson, 1981). It represents with emotional exhaustion, depersonalization, and loss of sense of accomplishment due to prolonged exposure to work-related stress (Agha et al., 2019; Biksegn et al., 2016; Leiter & Maslach, 2014; Moreira et al., 2018). In particularly, during residency period, stress is elongated in terms of time and intensity, and insufficient adaptation leads to burnout or psychobiological exhaustion of physicians (Ironside et al., 2019; Navinés et al., 2016). Burnout affects physicians’ clinical judgments, their ability to communicate and the provision of quality care for patients (Dyrbye et al., 2017; Elbarazi et al., 2017; Sanfilippo et al., 2017; Shah et al., 2019; Kane, 2019). In addition, burnout causes higher medical errors due to fatigue and reduced attention, which lead to neglect of personal safety precautions and patient safety (Dimitriu et al., 2020).

It is vital to address burnout in the context of the COVID-19 pandemic. In addition to the economic crisis and high unemployment rate, pandemic stress also affected the physical and psychological health of people (Blustein et al., 2020; Xiong et al., 2020). Countries confronted this period differently, so the impact of a pandemic on the quality of life varies according to the country and the individual (Suryavanshi et al., 2020). While the primary focus of frontline healthcare workers is to minimize transmission and treat patients with COVID-19, the impact, and consequences of the pandemic on mental health are significant (Fiest et al., 2021). During this period, mental health problems such as insomnia, anxiety, depression, burnout syndrome were observed in many study on healthcare workers (Barello et al., 2020; Brito-Marques et al., 2021; Pappa et al., 2020; Çevik & Ungan, 2021).

Several recent studies have found an increased level of burnout in healthcare professionals and medical assistants exposed to the pandemic. A study on medical residents during COVID-19 pandemic indicated a burnout prevalence of 76% (Osama et al., 2020) which is higher than the studies that are conducted before the pandemic (Malik et al., 2016; Serenari et al., 2019; Mion et al., 2013).

In the context of Türkiye, students graduate from medical school after 6 years as medical doctors. Who wants to get specialization chooses a department in a state or foundation owned university according to the score they got in the ‘Medical Specialization Exam’ (Mert et al., 2022; Sahin & Akcicek, 2005). Duration of medical residency training varies between 3-5 years, depending on the specialty area. In addition to theoretical training, residents take a central role in inpatient and outpatient care and night shifts (Küçükali & Akgöğ, 2021).

In 2020, there were a total of 26,181 residents in 68 university hospitals and 86 training and research hospitals in Türkiye. While there has been a significant transformation in many aspects of Türkiye’s health system in the last two decades, an adequate improvement has not been achieved in the work conditions of medical residents. Being overwhelmed by the heavy workload in the hospital, lack of time, energy and support for specialty training, low wages, mobbing, violence from patients and their relatives are among the ongoing problems (Küçükali & Akgöğ, 2021). According to the results of a survey conducted with 1069 medical residents from 11 provinces in Türkiye in 2007, the most common complaints during the residency training period are; excessive workload, excessive number of shifts and economic problems. In addition, medical residents stated that they found the training they received insufficient and that they had communication problems with their trainers (Aysan et al., 2008).

Although it is generally assumed that the incidence of burnout among physicians increased during the COVID-19 pandemic, we wanted to test this hypothesis on medical residents in a context where pre-pandemic level of burnout is already high. It was our chance to have a study (Küçükali et al., 2022) that was conducted in three university hospitals in Türkiye just before the pandemic. Using that study as a point of reference, the current study aims to compare the level of burnout in medical residents before and during the COVID-19 pandemic and identify potential risk factors.

## MATERIALS AND METHODS

This cross-sectional study is a replication of a previous study (Küçükali et al., 2022) which investigated the burnout level of medical residents in three university hospitals in Türkiye in December 2019, just before the COVID-19 pandemic emerged. The current study was conducted in the same hospitals in March 2021 (from March 5^th^ to May 5^th^, 2021) – one year after the pandemic hit Türkiye. There were approximately eight hundred medical residents in three hospitals combined. The study aimed to reach all active medical residents in hospitals excluding non-clinical departments (basic medical sciences, medical pathology, and public health).

Data was collected via a questionnaire that includes new questions regarding COVID-19 in addition to inherited the questions on sociodemographic and occupational factors, and Turkish adaptation of the Maslach Burnout Inventory (Ergin, 1992) from the previous study. Sociodemographic factors included age, sex, and marital status. Occupational factors included professional experience in years after obtaining medical degree, area of medical specialty, years spent in residency program, number of night shifts assigned last month. COVID-19 related factors include having a COVID-19 related duty, having infected by COVID-19 and living with someone at risk of severe COVID-19.

Well-established Maslach Burnout Inventory (MBI) measures burnout in 3 dimensions: emotional exhaustion (EE), depersonalization (DP), and lack of personal achievement (PA) via 22 questions. Questions used to be scored in 5-level Likert style ranging from “never” to “every day”. The inventory yields three scores from three sub-scales where burnout syndrome presented by high scores in emotional exhaustion and depersonalization, and low scores in personal accomplishment.

MBI is not a stand-alone diagnostic tool (Amosun & Dantile, 1996). Since there is no cut-off value, there is no distinction as having burnout or not (Amosun & Dantile, 1996; Pehlevan, 2017). The result is the probability of having burnout (Kleijweg et al., 2013). Although the three sub-dimensions of the burnout scale are related to each other, they are different concepts. For this reason, a total burnout score is not calculated during the evaluation (Pehlevan, 2017). While evaluating burnout in accordance with Maslach’s definition, the scores of each sub-dimension are collected and interpreted separately (Pehlevan, 2017; Williamson et al., 2018). In addition, as the burnout increases, the scores of the emotional exhaustion and depersonalization sub-dimensions increase, while the personal achievement score decreases (Pehlevan, 2017).

Researchers approached participants in their work environment (usually in doctors’ rooms in their clinics), informed them about the study, obtained their written consent and handed them a hard copy of the questionnaire. In the beginning, questionnaires were applied in a self-answered manner. However, researchers couldn’t reach all residents due to their unforseable shift schedules. For those who cannot be reached after two rounds of visits in all departments, we sent online versions of the questionnaires via instant messaging applications. 69 residents participated to study via online form.

To compare our results with the results of the aforementioned pre-pandemic study, we requested and obtained the data of the previous study. The combined dataset was analysed in Python (numpy v1.21.6, scipy v1.7.3, statsmodels v0.12.2). Mean total scores and standard deviations for each of the three subscales were calculated. Mean scores were compared between study periodsusing the independent samples t-test. Causal assumptions are presented in Figure 1. We build separate multiple linear regression models to estimate total effect of each variable on each of the three burnout sub-scale scores while avoiding “Table 2 Fallacy” (Westreich and Greenland, 2012). To eliminate possible confounding effects, we included confounder variables for given variable into equation according to Figure 1. Normality of distribution of the residuals are assessed with P-P plots. Statistical significance was assumed for the probabilities below 0.05 in statistical tests.This study was reviewed and approved by İstanbul Medipol University’s Non-invasive Clinical Studies Ethics Committee on 4.3.2021 (No: 284) and by the Ministry of Health on 5.3.2021.

**Figure 1.**
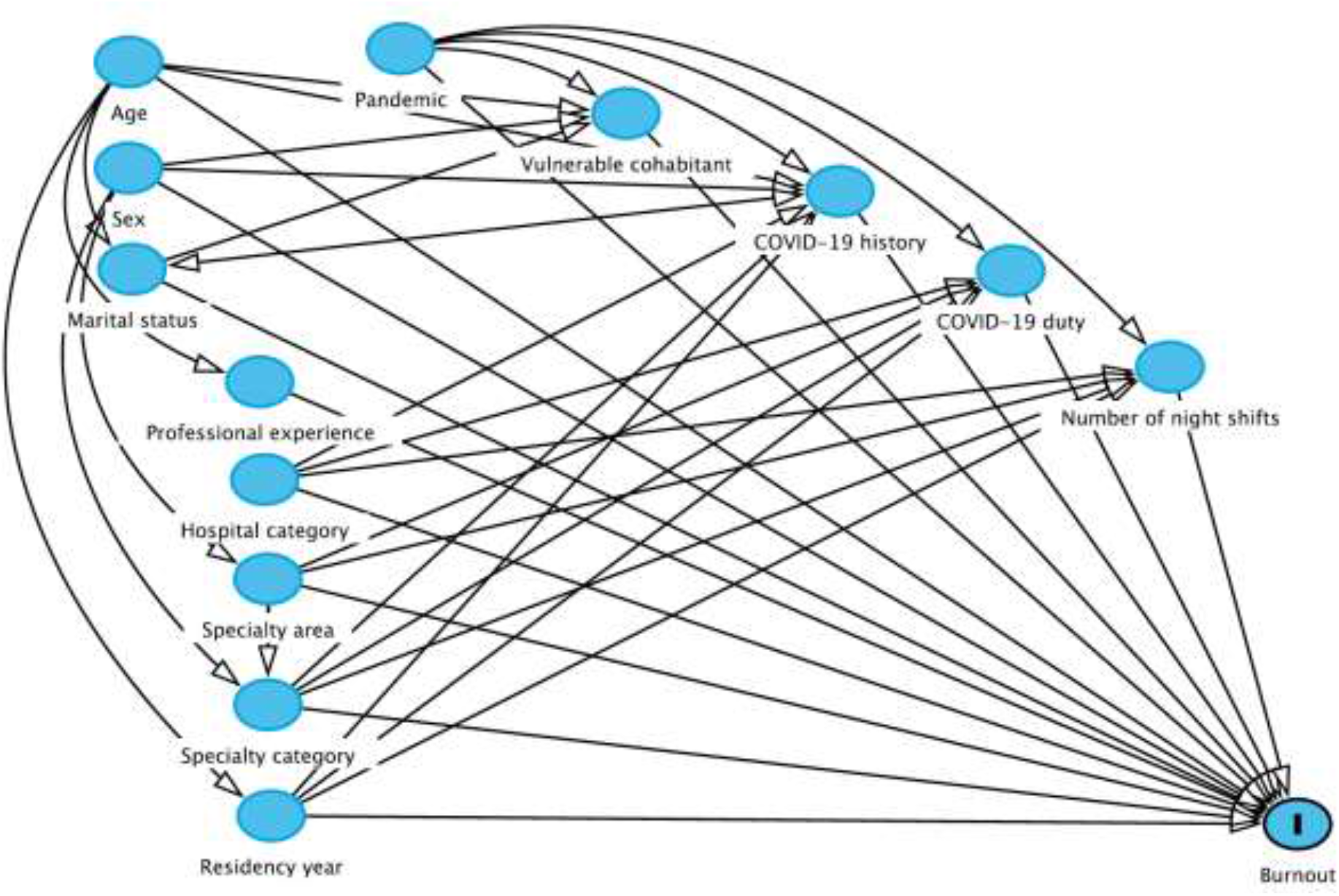
Directed acyclic graph showing causal assumptions.

## RESULTS

A total of 412 medical residents from three university hospitals participated in the current study. Participants’ characteristics including demographics and COVID-19 related variables were given alongside the previous study in Table 1. The mean age of the residents was 27.8±2.4 and half of them were female.

**Table 1.** Characteristics of the participants.

|  | Before the pandemic<br>(December 2019)* |  | During the pandemic<br>(March 2021) |  |
| --- | --- | --- | --- | --- |
|  | n | % | n | % |
| **Sex** |  |  |  |  |
| Female | 248 | 50.9 | 206 | 50.0 |
| Male | 239 | 49.1 | 206 | 50.0 |
| **Marital status** |  |  |  |  |
| Single | 310 | 63.7 | 294 | 71.4 |
| Married | 175 | 35.9 | 118 | 28.6 |
| Widowed | 1 | 0.2 | 0 | 0.0 |
| Divorced | 1 | 0.2 | 0 | 0.0 |
| **Hospital type** |  |  |  |  |
| Foundation owned | 196 | 40.2 | 258 | 62.6 |
| State owned | 291 | 59.8 | 154 | 37.4 |
| **Specialty group** |  |  |  |  |
| Internal | 291 | 59.8 | 265 | 64.3 |
| Surgical | 196 | 40.2 | 142 | 34.5 |
| Unknown | 0 | 0.0 | 5 | 1.2 |
| **Vulnerable cohabitant** |  |  |  |  |
| No | 487 | 100.0 | 341 | 82.8 |
| Yes | 0 | 0.0 | 71 | 17.2 |
| **COVID-19 diagnosis** |  |  |  |  |
| No | 487 | 100.0 | 267 | 64.8 |
| Yes | 0 | 0.0 | 145 | 35.2 |
| **COVID-19 duty** |  |  |  |  |
| Only Inpatient | 0 | 0.0 | 145 | 35.2 |
| Only outpatient | 0 | 0.0 | 32 | 7.8 |
| Both | 0 | 0.0 | 174 | 42.2 |
| None | 487 | 100.0 | 60 | 14.6 |
| Unknown | 0 | 0.0 | 1 | 0.2 |
| **Total** | 487 | 100.0 | 412 | 100.0 |
|  | **Mean** | **Std.** | **Mean** | **Std.** |
| **Age** | 27.9 | 2.4 | 27.8 | 2.4 |
| **Experience (years)** | 3.4 | 2.2 | 3.9 | 2.3 |
| **Residency year** | 2.3 | 1.1 | 2.2 | 1.3 |
| **Night shifts per month** | 6.0 | 2.9 | 6.2 | 2.7 |

\* Different from published paper (Küçükali et. al. 2022), participants from pathology department (n=3) are excluded in here.

The mean sub-scale scores of two studies compared in Table 2. Point estimates and distributions for each subscale score was similar in both study.

**Table 2.** Comparison of mean subscale scores before and during pandemic.

|  | Before the pandemic<br>(December 2019) |  | During the pandemic<br>(March 2021) |  | p |
| --- | --- | --- | --- | --- | --- |
|  | Mean | Std. | Mean | Std. |  |
| **Emotional Exhaustion** | 19.0 | 7.6 | 18.8 | 7.8 | 0.707 |
| **Depersonalization** | 7.3 | 4.3 | 7.2 | 4.4 | 0.792 |
| **Personal Accomplishment** | 20.8 | 5.1 | 21.1 | 5.0 | 0.310 |

Table 3 presents the total effect of each variable on burnout subscale scores. We built a separate linear regression model to estimate the total effect of each variable on each subscale score. In each model we adjusted for confounders specific to relationship of focus. Pandemic, i.e. study period, did not show a significant difference in any of the subscales. Emotional Exhaustion score is higher among who are female (1.77 [95%CI 0.76-2.77]), having trained in a surgical specialty (1.16 [95%CI 0.10-2.21]), have a vulnerable cohabitant (2.79 [95%CI 0.78-4.80]), and have more night shifts per month (0.46 [95%CI 0.25-0.66]). Depersonalisation score is higher among who spent more time in residency (0.29 [95%CI 0.00-0.58]), had a COVID-19 related outpatient duty (1.71 [95%CI 0.82-2.60]), and have more night shifts per month (0.23 [95%CI 0.12-0.35]). Personal Achievement score is lower among who are female (0.71 [95%CI 0.04-1.37]) and have a vulnerable cohabitant (1.66 [95%CI 0.28-3.03]).

**Table 3.**
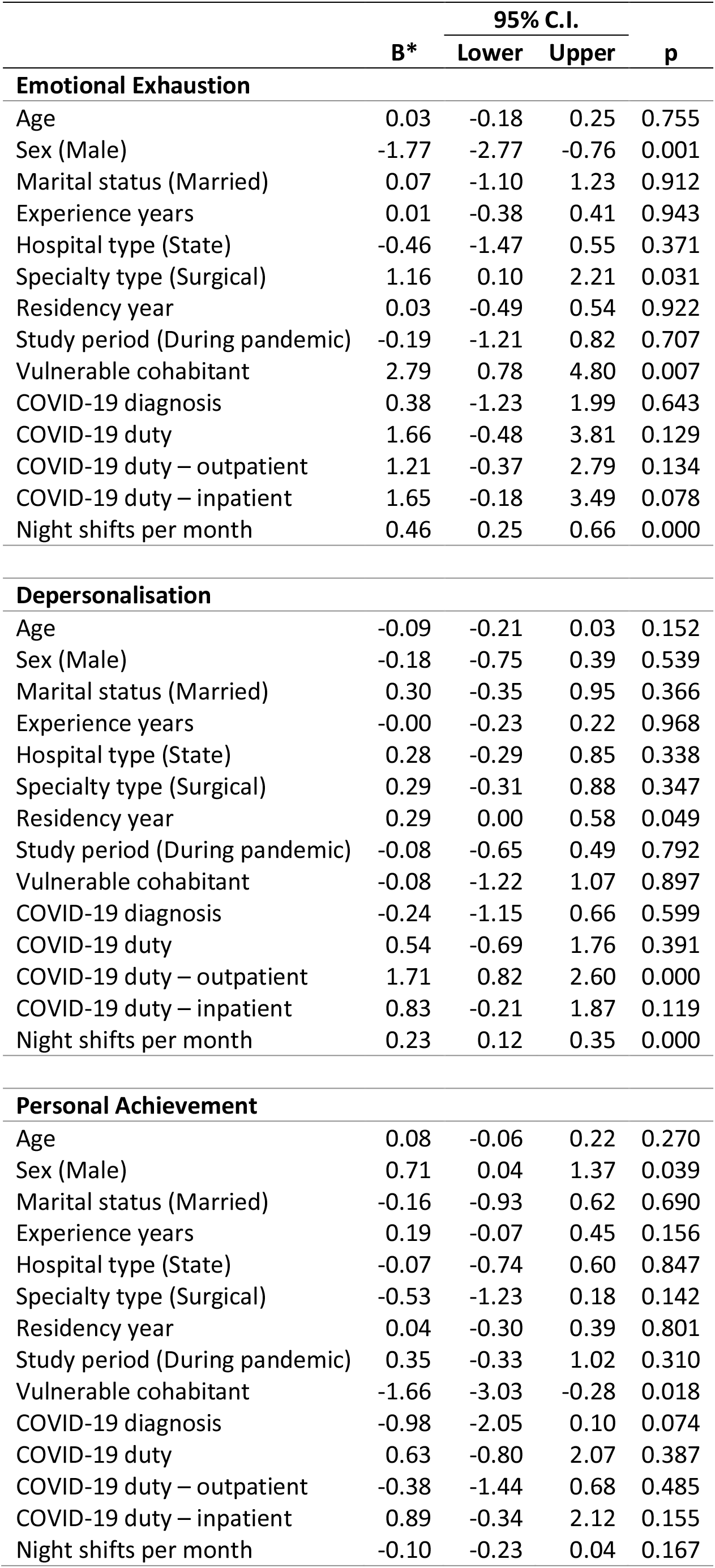
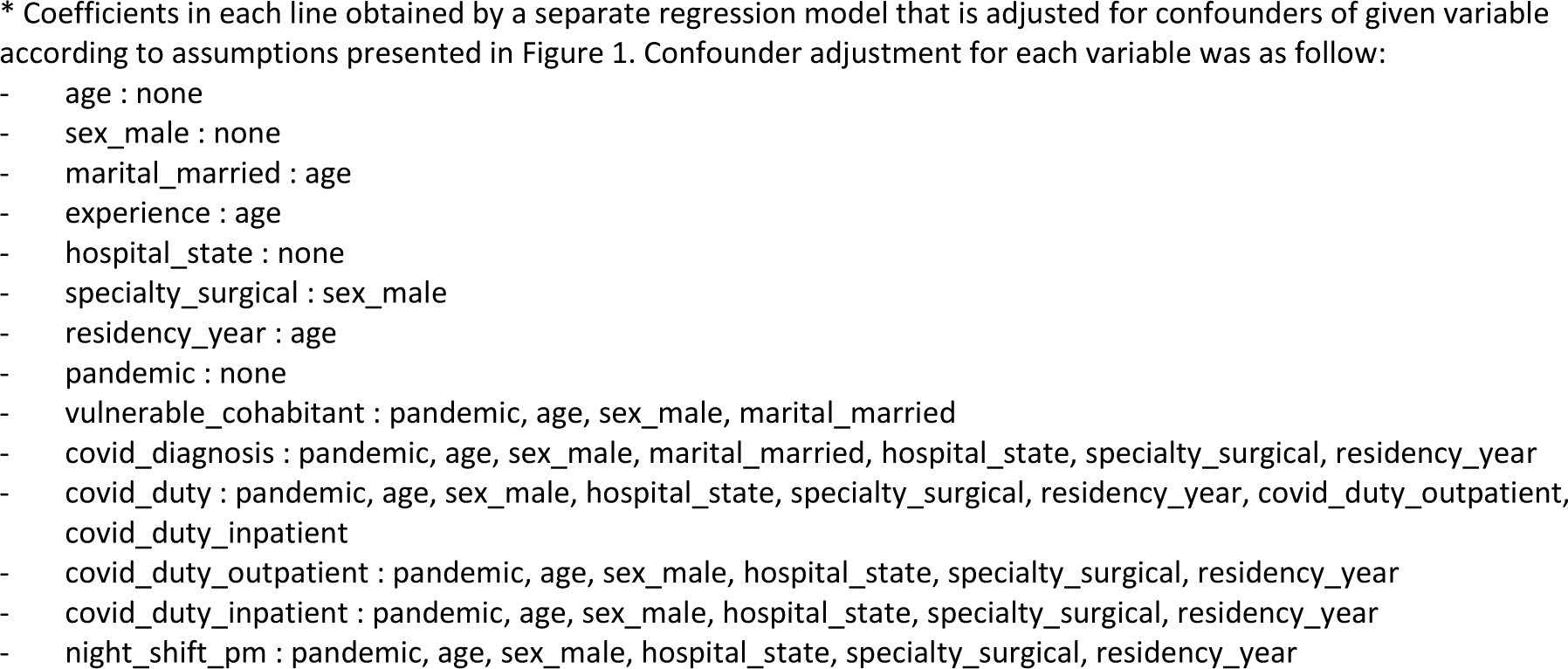
Total effect of each variable on burnout subscale scores.

|  | B* | 95% C.I. |  | p |
| --- | --- | --- | --- | --- |
|  |  | Lower | Upper |  |
| Emotional Exhaustion |  |  |  |  |
| Age | 0.03 | -0.18 | 0.25 | 0.755 |
| Sex (Male) | -1.77 | -2.77 | -0.76 | 0.001 |
| Marital status (Married) | 0.07 | -1.10 | 1.23 | 0.912 |
| Experience years | 0.01 | -0.38 | 0.41 | 0.943 |
| Hospital type (State) | -0.46 | -1.47 | 0.55 | 0.371 |
| Specialty type (Surgical) | 1.16 | 0.10 | 2.21 | 0.031 |
| Residency year | 0.03 | -0.49 | 0.54 | 0.922 |
| Study period (During pandemic) | -0.19 | -1.21 | 0.82 | 0.707 |
| Vulnerable cohabitant | 2.79 | 0.78 | 4.80 | 0.007 |
| COVID-19 diagnosis | 0.38 | -1.23 | 1.99 | 0.643 |
| COVID-19 duty | 1.66 | -0.48 | 3.81 | 0.129 |
| COVID-19 duty – outpatient | 1.21 | -0.37 | 2.79 | 0.134 |
| COVID-19 duty – inpatient | 1.65 | -0.18 | 3.49 | 0.078 |
| Night shifts per month | 0.46 | 0.25 | 0.66 | 0.000 |
| Depersonalisation |  |  |  |  |
| Age | -0.09 | -0.21 | 0.03 | 0.152 |
| Sex (Male) | -0.18 | -0.75 | 0.39 | 0.539 |
| Marital status (Married) | 0.30 | -0.35 | 0.95 | 0.366 |
| Experience years | -0.00 | -0.23 | 0.22 | 0.968 |
| Hospital type (State) | 0.28 | -0.29 | 0.85 | 0.338 |
| Specialty type (Surgical) | 0.29 | -0.31 | 0.88 | 0.347 |
| Residency year | 0.29 | 0.00 | 0.58 | 0.049 |
| Study period (During pandemic) | -0.08 | -0.65 | 0.49 | 0.792 |
| Vulnerable cohabitant | -0.08 | -1.22 | 1.07 | 0.897 |
| COVID-19 diagnosis | -0.24 | -1.15 | 0.66 | 0.599 |
| COVID-19 duty | 0.54 | -0.69 | 1.76 | 0.391 |
| COVID-19 duty – outpatient | 1.71 | 0.82 | 2.60 | 0.000 |
| COVID-19 duty – inpatient | 0.83 | -0.21 | 1.87 | 0.119 |
| Night shifts per month | 0.23 | 0.12 | 0.35 | 0.000 |
| Personal Achievement |  |  |  |  |
| Age | 0.08 | -0.06 | 0.22 | 0.270 |
| Sex (Male) | 0.71 | 0.04 | 1.37 | 0.039 |
| Marital status (Married) | -0.16 | -0.93 | 0.62 | 0.690 |
| Experience years | 0.19 | -0.07 | 0.45 | 0.156 |
| Hospital type (State) | -0.07 | -0.74 | 0.60 | 0.847 |
| Specialty type (Surgical) | -0.53 | -1.23 | 0.18 | 0.142 |
| Residency year | 0.04 | -0.30 | 0.39 | 0.801 |
| Study period (During pandemic) | 0.35 | -0.33 | 1.02 | 0.310 |
| Vulnerable cohabitant | -1.66 | -3.03 | -0.28 | 0.018 |
| COVID-19 diagnosis | -0.98 | -2.05 | 0.10 | 0.074 |
| COVID-19 duty | 0.63 | -0.80 | 2.07 | 0.387 |
| COVID-19 duty – outpatient | -0.38 | -1.44 | 0.68 | 0.485 |
| COVID-19 duty – inpatient | 0.89 | -0.34 | 2.12 | 0.155 |
| Night shifts per month | -0.10 | -0.23 | 0.04 | 0.167 |

\* Coefficients in each line obtained by a separate regression model that is adjusted for confounders of given variable according to assumptions presented in Figure 1. Confounder adjustment for each variable was as follow:
- age : none - sex\_male : none - marital\_married : age - experience : age - hospital\_state : none - specialty\_surgical : sex\_male - residency\_year : age - pandemic : none - vulnerable\_cohabitant : pandemic, age, sex\_male, marital\_married - covid\_diagnosis : pandemic, age, sex\_male, marital\_married, hospital\_state, specialty\_surgical, residency\_year - covid\_duty : pandemic, age, sex\_male, hospital\_state, specialty\_surgical, residency\_year, covid\_duty\_outpatient, covid\_duty\_inpatient - covid\_duty\_outpatient : pandemic, age, sex\_male, hospital\_state, specialty\_surgical, residency\_year - covid\_duty\_inpatient : pandemic, age, sex\_male, hospital\_state, specialty\_surgical, residency\_year - night\_shift\_pm : pandemic, age, sex\_male, hospital\_state, specialty\_surgical, residency\_year

## DISCUSSION

The main objectives of our study were to compare the level of burnout in medical residents before and during the COVID-19 pandemic and identify risk factors that may be associated with burnout. Our findings suggest that during pandemic medical residents have similar but high levels of burnout as before the pandemic. We have identified that having a vulnerable cohabitant and outpatient COVID-19 duty are risk factors for burnout during pandemic alongside known factors such as female sex, surgical specialty, residency year and number of night shifts.

In the literature a range of causes is suggested for increased burnout during the pandemic. New tasks due to the pandemic, fear of illness, lack of knowledge on using personal protective equipment (PPE), inadequate PPE, faulty infection control measures, faulty communication and directives, lack of emotional support and preparation, low autonomy, lack of management support, low hospital management support, lack of appreciation from supervisors, job insecurity, perceived fatality, not being able to spare enough time for the patient, fear of mistakes, medical neglect, dissatisfaction with patient care, fear of carrying a disease to the family, staying in a place other than the family home for security measures, not being able to see the family, being young, isolation and stigma are some of those(Aba, 2022; Dumea et al., 2022; Kılıç et al., 2021; Sengul et al., 2021; Soares et al., 2022).

Seda-Gombau et. al. conducted series of cross-sectional studies in November 2016, January 2019, and October 2020 to see the change of burnout levels among primary care physicians in Catalonia. Although the study has quite small sample size, it showed increase in burnout as a result of upward trend in EE and DP, and downward trend in PA subscales by the pandemic (Seda-Gombau et al., 2021). Contrary to that study and our expectations, our study does not support the hypothesis of higher burnout during the pandemic. This may be due to already high levels of burnout among medical residents in Türkiye. Another explanation would be that exacerbating effects of the pandemic may be balanced by the relative decrease in demand of other (elective) health services and imposed flexible work schedules during pandemic. Anecdotally, some of our participants commented that except COVID-19 specific duties, their overall workload is actually decreased.

On the other hand, our findings regarding burnout subscale scores are consistent with many studies that are conducted in different regions of Türkiye throughout the pandemic. For example, in April 2020 Kılıç et. al. measured burnout levels of 748 physicians from 59 cities of Türkiye via online convenience survey where EE was 19.7 ± 8, DP was 6.9 ± 4.5 and, PA was 8.9 ± 5.1 (Kılıç et al., 2021). It is worth noting that this study reverse-coded PA subscale and if our study was coded same way our estimation of PA score would be 10.9 ± 5.0. In May 2020 Sevinç et. al. estimated median EE as 22 (IQR: 15-28.7), DP as 6 (IQR: 3-9), and PA as 23 (IQR: 17-26) among 104 physicians, residents, and nurses in an intensive care unit in Istanbul (Sevinc et al., 2022). Similarly, in December 2020, Aba estimated mean EE as 20.3 ± 8.8, DP as 7.9 ± 4.4, and PA as 20.5 ± 4.6 among 350 healthcare workers in Türkiye (Aba, 2022). Those studies provide not only similar point estimates but also similar dispersion for three subscale scores. Of course, heterogeneity in some degree between regions and professional should be expected.

It is understandable that emotional exhaustion, which is evaluated as feeling emotionally distant from work, feeling tired at the beginning and end of the workday, feeling nervous, stressed and overworked due to working with people all day, exhausted and disappointed, helpless due to work, is high in healthcare workers living with a person at risk of COVID-19.

In studies, EE scores were found to be higher in women, single people, those with less professional years, with longer working hours, those who experienced sleep deprivation, and who worked in shifts (Kılıç et al., 2021; Seda-Gombau et al., 2021; Sengul et al., 2021). The effects of work-home conflicts and maternal responsibilities on female health workers are known (Kılıç et al., 2021; Soares et al., 2022). In our study, multiple regression analysis showed that woman have higher the EE score. It is known that the high number of night shifts that cause sleep deprivation and poor-quality sleep which increases burnout (Seda-Gombau et al., 2021; Sevinc et al., 2022). In our study, multiple regression analysis also showed that higher number of night shifts per month is associated with higher the EE score.

Again, in some studies conducted during the pandemic period, the emotional exhaustion scores of healthcare workers who were in direct contact with patients due to the high infectivity of COVID-19, and who were afraid of infecting their family members with COVID-19 were found to be high (Jose et al., 2020; Liu et al., 2020; Luceño-Moreno et al., 2020; Serrão et al., 2021; Soto-Cámara et al., 2021).

In our study, analysis showed that more time spent in residencyand higher number of night shifts per month is associated with higher DP score.

Some studies show that the residency year does not make a difference in DP (Passos et al., 2022; Wilson et al., 2017). It is known that exposure to criticism and violence increases DP scores (Passos et al., 2022; Wilson et al., 2017). In our study, the fact that the DP score was high in the younger ones but the increase in residency year was a positive predictor of DP may be evaluated in this context. It was found that the burnout levels (EE, DP, and PA) of night shift workers were significantly higher than those working the day shift (Hacimusalar et al., 2021; Sengul et al., 2021). In our study, it was reported that DP scores were higher in shift workers, which was consistent with the literature.

In the early period of the pandemic, that is, at a time when there are many uncertainties, the burnout frequency of healthcare professionals working on the front line was found to be lower. (Wu et al., 2020). However in our study, residents who worked in COVID-19 outpatient services has the higher depersonalisation score. This difference can be related to duration of the exposure and prolonged exposure may lead to depersonalisation.

Different relationships were found between gender and PA (Aba, 2022; Sengul et al., 2021). Although in some studies, as ours, men had a higher average of PA than women, in other studies, it was the opposite (Aba, 2022; Kılıç et al., 2021; Sengul et al., 2021). In a study examining the causal ranking of the three sub-dimensions of burnout among male and female physicians in 2011; It was found that personal accomplishment for men developed independently of the other two sub-dimensions. Despite an increase in emotional exhaustion and depersonalization, a sense of personal accomplishment has increased among male physicians. For men, it was thought that the decrease in personal accomplishment was not a dimension of burnout and they had a sense of self-efficacy that was not affected by emotional exhaustion and depersonalization. For women, it was observed that the sense of personal accomplishment was affected by the other two sub-dimensions of burnout. It has been stated that women who experience emotional exhaustion and become desensitized to their patients may feel guilty and think that they are personally unsuccessful(Houkes et al., 2011).

COVID-19 is, undoubtedly, the biggest distruption that health systems have faced in last century. It is fair to expect it to increase stress and, by extension, burnout among healthcare workers. However, our study does not support this hypothesis. We could not find a significant difference burnout levels of medical residents before and during pandemic. Could it simply be explained by selection bias or small sample size? Despite many others our study did not used convenience sampling with online surveys and tried to reach most of the residents in investigated hospitals. We were able to reach approximately half of the population and distribution of participants were quite similar with reference study. Regarding the medical residents, to our knowledge, we included more participants than any other studies conducted during pandemic in Türkiye.

We would like to raise four possible explanations and hypotheses for further research. First, burnout levels were already high before pandemic and pandemic did not / could not increase them further. Second, in contrary to induced demand beforehand, decrease in demand to other (elective) health services and relatively flexible work schedules during pandemic created a space where medical residents can relax. Third, capacity of health system in Türkiye was able to manage stress on the system. Fourth, pandemic management was successful to protect health system from excess workload. Nevertheless, medical residents are facing serious levels of burnout. As central players in the health system their problems should be always a priority in health management until we reach better working conditions for medical residents.

## Limitations

It is important to note that this study was not longitudinal, i.e., data is not necessarily collected from the same residents in the reference study. There is a constant change in residents in each hospital. Thus, this study does not show a causal relationship between the pandemic and burnout among medical residents. Still, the comparison in this study provides better evidence than existing studies in the Turkish context for both local decision-makers and global researchers. Some may argue that multiple comparisons require adjustment for inflation in familywise type 1 error. We did not see a need for adjustment for multiple comparisons. Because we only tested familywise hypotheses while we were comparing burnout levels of two groups and none of the three was significant even at 0.05 level (Table 2). Although the study has achieved an approximate 50% participation rate (exact number of all residents in the hospitals was not provided to us), selection bias should be always kept in mind while interpreting the findings. Residents who experience burnout may be more or less willing to participate in the study than others. For example, percentage of residents from state owned hospital was lower in the current study than previous study. Their participation may be related with their burnout. Lastly, 69 participants answered the online form, and they have a significantly higher mean EE score (21.9±7.5) than those who answered the paper form (18.2±7.7).

## Data Availability

Data used in this study is available on request from the corresponding author.

## Acknowledgements

Authors would like to thank our interns Abidin Fatih Emhan, Ahmet Burak Börekçi, Kadir Kolçak, and Serdar Yıldız for their support in data collection.

## Ethical approval

This study is reviewed and approved by İstanbul Medipol University’s Non-invasive Clinical Studies Ethics Committee on 4.3.2021 (No: 284) and by the Ministry of Health on 5.3.2021.

## Data availability

Data used in this study is available on request from the corresponding author.

